# The Kids Are Not Alright: A Preliminary Report of Post-COVID Syndrome in University Students

**DOI:** 10.1101/2020.11.24.20238261

**Authors:** Julie Walsh-Messinger, Hannah Manis, Alison Vrabec, Jenna Sizemore, Karyn Bishof, Marcella Debidda, Dolores Malaspina, Noah Greenspan

## Abstract

**Background:** Post-COVID syndrome is increasingly recognized by the medical community but has not been studied exclusively in young adults. This preliminary report investigates the prevalence and features of protracted symptoms in non-hospitalized university students who experienced mild-to-moderate acute illness.

**Methods:** 148 students completed an online study to earn research credit for class. Data from COVID-19 positive participants with symptoms ≥28 days (N=22) were compared to those who fully recovered (N=21) and those not diagnosed with COVID-19 (N=58).

**Results:** 51% of participants who contracted COVID-19 (N=43) experienced symptoms ≥28 days and were classified as having post-COVID syndrome; all but one (96%) were female. During acute illness the post-COVID group, compared to those who fully recovered, experienced significantly more chest pain (64% vs 14%; P=.002), fatigue (86% vs 48%; P=.009), fever (82% vs 48%; P=.02), olfactory impairment (82% vs 52%; P=.04), headaches (32% vs 5%; P<.05), and diarrhea (32% vs 5%; P<.05). Compared to those not diagnosed with COVID-19, the post-COVID syndrome group more frequently experienced exercise intolerance (43% vs. 0%; P<.001), dyspnea (43% vs. 0%; P<.001), chest pain (31% vs 7%; P=.002), olfactory impairment (19% vs 0%; P=.004), lymphadenopathy (19% vs 0%; P=.004), gustatory impairment (14% vs 0%; P=.02), and appetite loss (36% vs 14%; P=.02).

**Interpretation:** Our results contradict the perception that this “yet to be defined” post-COVID syndrome predominantly affects middle-aged adults and suggest that exercise intolerance, dyspnea, chest pain, chemosensory impairment, lymphadenopathy, rhinitis, and appetite loss may differentiate post-COVID syndrome from general symptoms of pandemic, age, and academic related stress. These findings are also consistent with previous reports that females are more vulnerable to this post viral syndrome. Large-scale population-based studies are essential to discerning the magnitude and characterization of post-COVID syndrome in young adults as well as more diverse populations.

## INTRODUCTION

The novel SARS-CoV-2 virus, first reported in Wuhan in late 2019, is an international pandemic that has infected over 58 million and resulted in over 1.3 million deaths as of November 23, 2020. The initial expectation that mild and moderate COVID-19, the disease caused by SARS-CoV-2, would resolve within 2-3 weeks has been challenged by anecdotal reports of prolonged and debilitating symptoms in COVID-19 survivors.^1-3^ Studies, such as Carfì et al’s Italian case series which found that 87.4% of 143 cases who were hospitalized for COVID-19 continued to experience symptoms two months after their positive test result,^4^ have resulted in increasing recognition of prolonged symptoms and disability for adults who were hospitalized with severe COVID-19.

Young adults are widely considered to have a low risk of significant COVID-19 symptoms, but newer studies estimate that many infected adolescents and young adults may require hospitalization.^5,6^ The notion that COVID-19 has a minimal impact on the health of young adults is contrary to a CDC report which found one in five individuals without pre-existing medical conditions who were between 18-34 years old contracted the disease reported via telephone contact that they were still symptomatic three weeks after their positive test.^7^ This assertion is further contradicted by reports in September and October, 2020 of three healthy male university student fatalities directly related to COVID-19, two of whom were athletes.^8^ As there are currently no reports of protracted symptoms in non-hospitalized young adults who contracted COVID-19, this preliminary report investigates the prevalence and features of post-COVID syndrome in a sample of university students with mild to moderate acute illness severity.

## METHODS

### Participants

Undergraduate students (N=195) at a private, Catholic, Midwestern university completed an online study about COVID-19 testing, symptoms, course of illness, treatment, and current functioning between October 7 and November 11, 2020. Participants were recruited through SONA, an online research study management system, and the self-report measures were administered through Qualtrics (Qualtrics^XM^, Provo, UT). The study was approved by the University of Dayton Research Review and Ethics Committee, a subcommittee of the Institutional Review Board, and participants provided voluntary informed consent obtained in accordance with the Declaration of Helsinki. Participants were compensated for their time with research experience credit for class.

### Measures

A health survey, developed for this study by JWM and KB, asked about demographics, medical history, COVID-19 testing and results, course of illness, treatment, and past and current COVID-19 symptoms, including fever, pharyngitis, lymphadenopathy, cough, dyspnea, rhinitis, myalgias, fatigue, diarrhea, constipation, appetite loss, nausea, emesis (excluding alcohol related incidents), headache, and chemosensory impairment. The survey also asked about post-COVID onset of exercise intolerance, hypertension, hypotension, tachycardia, bradycardia, brain fog, and impaired concentration, memory, and sleep. Overall severity of illness and treatment were assessed with questions 2a, 2b, and 2c from the *COVID Experiences (COVEX) Symptoms and Diagnoses* module.^9^

Perceived stress over the past month was assessed with the *Perceived Stress Scale* (PSS).^10^ The 10-items on this self-report measure are rated on a 5-point Likert scale from “never” to “very often” and are summed to yield a total score that can range from 0 to 40.

Depression severity over the past two weeks was measured with the *Center for Epidemiological Studies Depression Scale – Revised* (CESD-R).^11^ The 20-item self-report questionnaire produces an overall severity score that ranges from 0 to 60, as well as subscale scores for Dysphoria and Sleep (subscales range from 0-9), Anhedonia, Appetite, Thinking/Concentration, Guilt, Fatigue, Agitation, and Suicidal Ideation (subscales range from 0-6).

The *Generalized Anxiety Disorder-7* (GAD-7)^12^ was used to assess generalized anxiety severity over the past two weeks. The seven self-report items are rated on a 4-point scale from “not at all” to “nearly every day” and total scores can range from 0 to 28.

### Main Outcome Variables

The primary outcomes measures were COVID-19 positivity and duration of symptoms. Secondary outcomes included treatment utilization, depression, anxiety, and perceived stress.

### Statistical analysis

SPSS (version 27.0) was used for data analysis, which was conducted by JWM. Descriptive statistics of all measures were examined to identify key features of the data (e.g., non-normal distribution, outliers, skewness) that might influence inferential methods. All variables were determined to be normally distributed, with the exception of age days since illness onset, and depression severity, which were positively skewed.

Differences between the participants who reported full recovery within 28 days (N=21) and those with post-COVID syndrome (defined as protracted symptoms ≥28 days; N=22) were assessed with independent samples t-tests for normally distributed continuous variables or Mann Whitney U-tests for non-normally distributed continuous variables that were not. Cohen’s d, Common Language Effect Size (CLES) and 95% confidence intervals [CI] were computed using the Psychometrica online calculator.^12^ Chi-Square and Fisher’s Exact tests were used to examine group differences in categorical variables, the latter employed when cell counts were <5, with Cramer’s V computed for effect sizes. Reported P-values are all two-tailed.

### Role of the Funding Source

The funder had no role in the study design, data collection, analysis, interpretation, writing of this manuscript or decision to submit to The Lancet for publication. JWM, HM, AV, and JS had access to all of the data. The corresponding author, JWM, had final responsibility for the decision to submit the manuscript for publication, to which all authors agreed.

## RESULTS

Of the 215 students who enrolled in the study, 93% completed the self-report measures (N=195). Excluded from analyses were 27 students who had no COVID-19 testing or diagnosis, seven who did not know their test results, four with missing data, and nine who were <28 days post onset of COVID-19 at the time they completed the measures, yielding a final sample size of 148 (see Figure 1). The university requirement that students produce a negative COVID-19 test before returning to campus for the Fall 2020 semester was the most frequent reason for COVID-19 testing (38%), followed by COVID-19 symptoms (26%), close contact with a known positive case (23%), random testing (7%), and other reasons (6%), which included peace of mind and requirement prior to a medical procedure.

**Figure 1.**
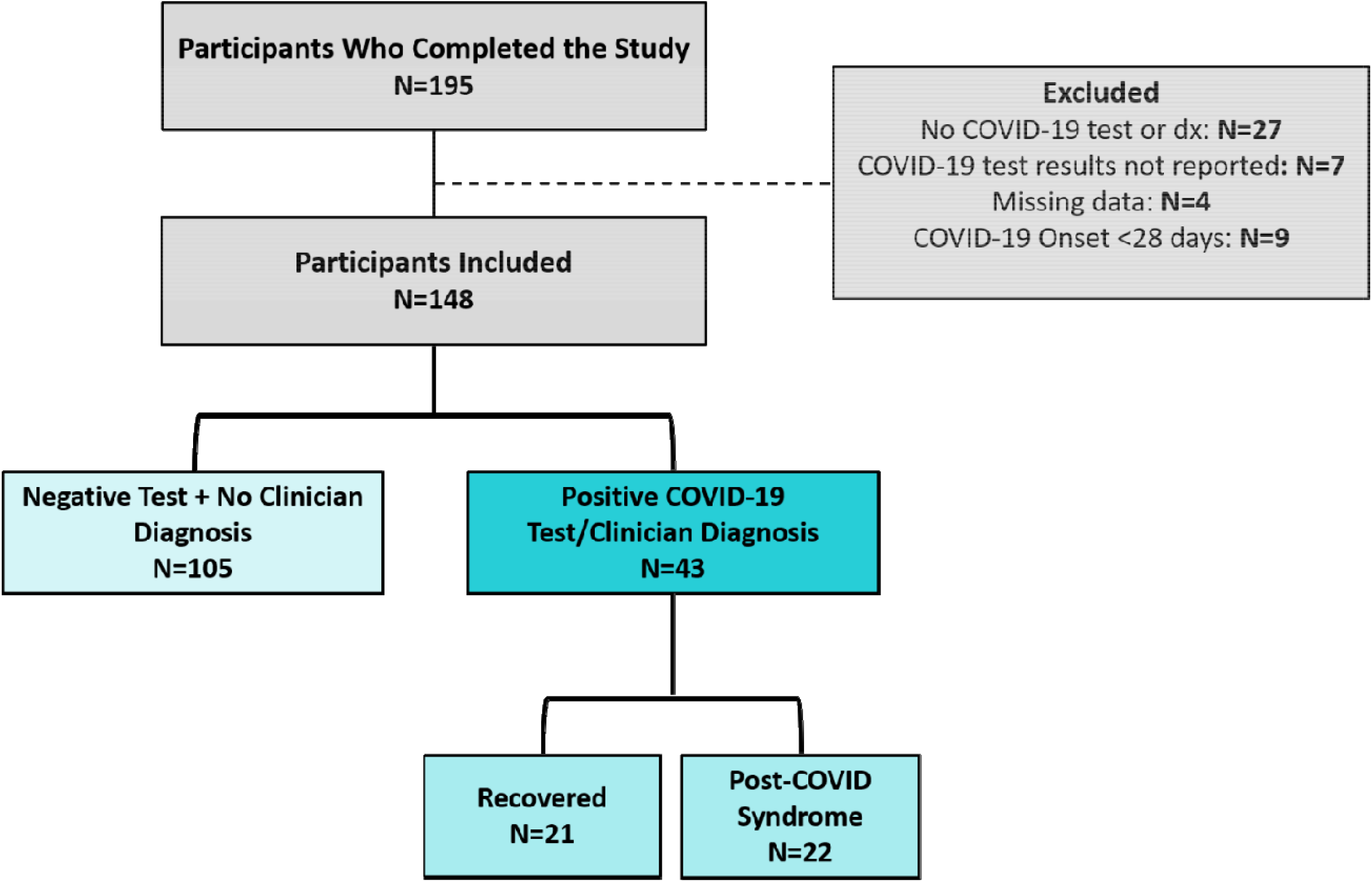
Breakdown of included and excluded study participants

One third of the sample (N=52) reported a positive COVID-19 test result or diagnosis. Participants primarily had their diagnosis confirmed by PCR testing (76%); however, seven (16%) were unable to obtain testing but were diagnosed by a clinician, and four (9%) had a negative test but were nonetheless diagnosed as having COVID-19 by a clinician in light of smell loss and other symptoms. Means, standard deviations, and frequency counts for all demographic measures are presented in Table 1.

**Table 1.** Demographic characteristics of the COVID negative, COVID positive recovered, and post-COVID syndrome groups.

|  | **COVID**<br>**Negative<sup>†</sup>**<br>N=58 | **COVID**<br>**Recovered**<br>N=21 | **Post-COVID**<br>**Syndrome**<br>N=22 |
| --- | --- | --- | --- |
| Age, years — M ± SD | 18.91 ± 1.04 | 18.95 ± 0.74 | 19.86 ± 3.03 |
| Sex — n (%) |  |  |  |
| Male | 21 (36.2) | 7 (33.3) | 1 (4.5) |
| Female | 37 (63.8) | 14 (66.7) | 21 (95.5) |
| Race — n (%) |  |  |  |
| Non-White | 6 (10.3) | 0 (0.00) | 1 (4.5) |
| White | 52 (89.7) | 21 (100) | 21 (95.5) |
| Ethnicity — n (%) |  |  |  |
| Hispanic | 5 (8.6) | 1 (4.8) | 1 (4.5) |
| Not Hispanic | 53 (91.4) | 20 (95.2) | 21 (95.5) |
| Cigarette Use — n (%) |  |  |  |
| None | 52 (89.7) | 21 (100.0) | 18 (81.08) |
| Yes, but not daily | 6 (10.3) | 0 (0.0) | 4 (18.2) |
| Use of Vaping Devices — n (%) |  |  |  |
| None | 39 (67.2) | 14 (66.7) | 16 (72.7) |
| Yes, but not daily | 14 (22.1) | 6 (28.8) | 5 (22.7) |
| Yes, daily | 5 (8.6) | 1 (4.8) | 1 (4.5) |

<sup>†</sup>Demographic data are only reported for participants for which current symptom data was available.

**Table 2.** Characteristics of acute COVID-19 experienced by the COVID positive recovered and post-COVID syndrome groups.

|  | **COVID**<br>**Recovered**<br>**N=21** | **Post-COVID**<br>**Syndrome**<br>**N=22** |
| --- | --- | --- |
| Days post COVID-19 onset, M ± SD | 93.50 ± 72.75 | 79.32± 70.23 |
| Number of COVID-19 symptoms during acute illness, M ± SD | 5.24 ± 2.91 | 9.09 ± 3.91 |
| Overall COVID-19 severity — n (%) |  |  |
| Mild | 16 (76.2) | 8 (36.4) |
| Moderate | 4 (19) | 13 (59.1) |
| Severe | 1 (4.8) | 1 (4.5) |
| Treatment — n (%) |  |  |
| Saw healthcare provider in person (e.g. clinic, doctor's office, university health center, urgent care, or emergency room) | 2 (9.5) | 6 (27.4) |
| Spoke to a healthcare provider over the phone, by email, or online | 4 (19.0) | 8 (36.4) |
| No contact with a healthcare provider | 14 (71.4) | 8 (36.4) |

### Post-COVID syndrome participants compared to recovered participants

#### Demographics

While 91% of COVID-19 positive participants reported “full recovery” from acute illness, 51% (N=22) continued to experience protracted symptoms, of whom 59% (N=13) had persistent symptoms for ≥50 days. The number of days since the onset of COVID-19 ranged from 28 to 291 and the groups did not differ (U=171.00; Z=-1.46; P=.15; CLES=0.56; CI, -0.40 to 0.80). The recovered group reported an average of seven days from onset to symptom remission. There were significantly more females in the post-COVID syndrome group compared to the recovered group (*Χ*^2^=5.88, P=.02, Cramer’s V=0.37). The groups did not differ with regard to age, race, ethnicity, cigarette smoking, or vaping (all P’s≥.11).

The rates of pre-existing chronic illnesses were low across both groups. The most frequently reported chronic illness was allergic rhinitis, which was endorsed by 29% of recovered participants and 36% of the post-COVID syndrome group. One participant in the post-COVID group reported a history of hypertension and irritable bowel syndrome, one had endometriosis and fibromyalgia, and three others reported asthma. One recovered participant reported obesity, two endorsed asthma, and one had a rare connective tissue disorder.

#### Symptomatology and severity

One participant in each group reported a severe course of illness (e.g. severe dyspnea or complications leading to pneumonia). Even with these cases excluded, the post-COVID syndrome group more frequently reported moderate illness severity compared to the majority of the recovered group who were asymptomatic (N=1) or reported a mild course (P=.01; Cramer’s V=0.43).

During acute illness, the post-COVID syndrome group, compared to the recovered group, experienced significantly more chest pain chest pain (P=.002; Cramer’s V=0.51), fatigue (*Χ*^2^=6.86; P=.009; Cramer’s V=0.40), fever (*Χ*^2^=5.32; P=.02; Cramer’s V=0.40), olfactory impairment (*Χ*^2^=4.24; P=.04; Cramer’s V=0.31), headaches (P<.05; Cramer’s V=0.35), and diarrhea (P<.05; Cramer’s V=0.348; see Figure 2). No group differences were observed in the frequency of other symptoms (all P’s>.14). The number of symptoms experienced during acute illness was significantly higher in the post-COVID syndrome group (t=-2.77; P=.008; d=0.82; CI, 0.24 to 1.42) who were also more likely to have sought medical treatment (*Χ*^2^=9.79; P=.002; Cramer’s V=0.47).

**Figure 2.**
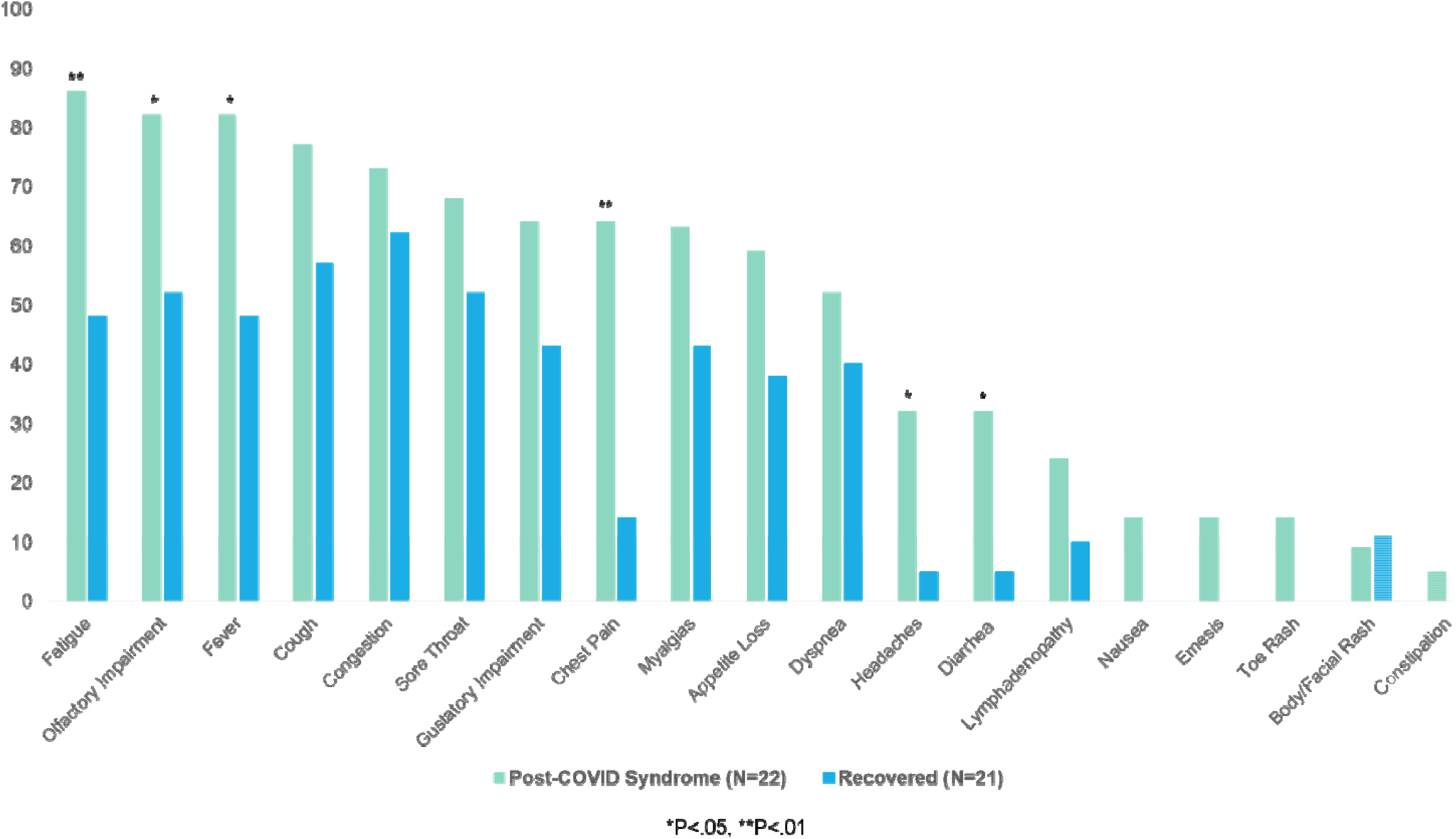
Symptoms reported by COVID recovered and post-COVID syndrome participants during acute illness. Bars represent the percentage of each group who endorsed each symptom.

Depression severity was higher in the post-COVID syndrome group (U=148.00; Z=-2.02; P=.04; CLES=0.58; CI, -0.31 to 0.90); however, the only depression subscales that differed were Sleep (U=145.5; Z=-2.11; P=.04; CLES=0.61; CI, -0.21 to 0.99), and Thinking (U=146.00; Z=-2.41; P=.02; CLES=0.64; CI, -0.12 to 1.09), with a marginal difference for Appetite (U=157.00; Z=-1.93; P=.05; CLES=0.66; CI, -0.02 to 1.21). Perceived stress and anxiety also did not differ between the groups (P’s ≥.47).

### Characterization of participants with post-COVID syndrome

The number of symptoms and/or complications experienced by the post-COVID syndrome group in the past week ranged from 2 to 33 (*M*=7.00±7.60), with mean symptom severity (rated on a 5-point scale from “mild” to “severe”) ranging from 1.0 - 3.21 (*M*=1.74± 0.78). Impaired concentration was the most frequently reported symptom, followed by headache, rhinitis, exercise intolerance, dyspnea, sleep impairment, brain fog, appetite loss, fatigue, and chest pain. None of the study participants were hospitalized; however, six (27%) reported in-person contact with at least one healthcare provider (e.g., primary care physician, university health center, urgent care, or emergency room) and eight others (36%) reported telehealth contact. Despite experiencing serious symptoms including chest pain, dyspnea, and exercise intolerance, eight participants (36%) reported no contact with any healthcare professional.

### Comparison of Post-COVID syndrome to COVID negative participants

As some of the current symptoms endorsed by the post-COVID syndrome group could be attributed to allergic rhinitis or other viral infections, we compared the frequency of their symptoms to those endorsed by COVID negative participants for whom current symptom data was available (N=58) and who served as an ecologically valid normative sample. The post-COVID syndrome group more frequently reported exercise intolerance (P<.001; Cramer’s V=0.60), dyspnea (P<.001; Cramer’s V=0.60), chest pain (P=.002; Cramer’s V=0.38), olfactory impairment (P=.005; Cramer’s V=0.38), lymphadenopathy (P=.004; Cramer’s V=0.37), rhinitis (*Χ*^2^=8.57; P=.003; Cramer’s V=0.33) and gustatory impairment (P=.017; Cramer’s V=0.33) and appetite loss (*Χ*^2^=5.08; P=.02; Cramer’s V=0.25; Figure 3). Notably, one or more of the eight aforementioned symptoms were endorsed by all but one post-COVID syndrome participant (M=1.91±0.41).

**Figure 3.**
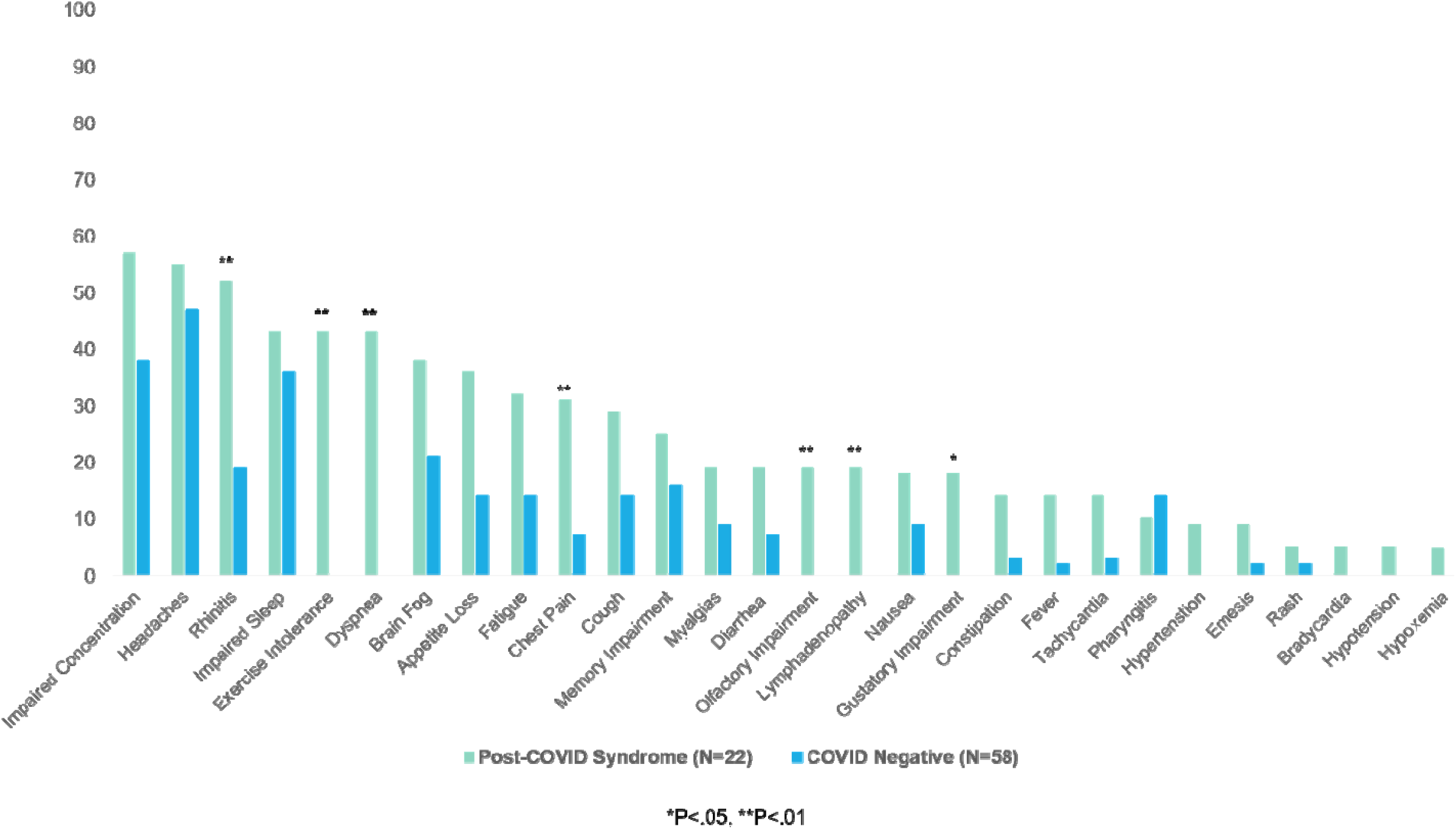
Symptoms endorsed over the past week by post-COVID syndrome participants and those who tested negative for COVID-19. Bars represent the percentage of those who endorsed the symptom in each group.

While the results did not reach statistical significance, fever (P=.06; Cramer’s V=0.24), fatigue (*Χ*^2^=3.40; P=.07; Cramer’s V=0.21), and hypertension (P=0.07; Cramer’s V=0.26) tended to also be more frequent in the post-COVID syndrome group with moderate effect sizes. There were no differences in frequency of pharyngitis, myalgias, diarrhea, constipation, hypotension, tachycardia, bradycardia, brain fog, and sleep, memory, and concentration impairment (all P’s>.10). Post-COVID syndrome participants also endorsed a greater number of current symptoms (U=315.00; Z=-3.54; P<.001; CLES=0.71; CI, 0.16 to 1.36) and greater mean severity of current symptoms (U=375.00; Z=-2.88; P=.004; CLES=0.53; CI, -0.48 to 0.72) compared to COVID negative participants. Severity of depression, anxiety, and perceived stress did not significantly differ between the groups (all P’s ≥.200).

## Discussion

The importance of understanding the COVID-19 disease in young adults cannot be overstated, as the weekly number of such cases reported to the CDC is increasing over time and especially as students have returned to campus settings.^13^ This preliminary study contradicts the perception that this “yet to be defined” post-COVID syndrome predominantly affects middle-aged adults, as just over half of our young adult sample reported at least two COVID-19 symptoms and/or related complications 30 days post illness onset, at minimum; a majority were still symptomatic more than 50 days later. Our comparison of post-COVID syndrome participants to those never diagnosed with COVID-19 suggests that the protracted symptoms experienced by the post-COVID syndrome group are not merely due to stress associated with the current pandemic or college attendance.

Of paramount concern is that a third of the post-COVID syndrome group had not had contact with a healthcare provider, despite serious symptoms, including dyspnea, exercise intolerance, chest pain, and chemosensory impairment. The current message in the United States that COVID-19 is benign and/or a short lived illness in young adults is inconsistent with reports that adolescents and young adults may require hospitalization,^5,6^ and several recent fatalities in healthy university students.^8^

Understandably, scientific efforts since the outbreak of SARS-CoV-2 have predominantly focused on prevention (e.g., vaccine development, studying the efficacy of social distancing and masks, developing early detection tests) and development of treatment approaches to reduce the morbidity and mortality associated with COVID-19, with little attention within the scientific and medical community to the lingering effects of the disease. As a result, individuals with protracted COVID symptoms, frequently referred to as “long-haulers” or “long COVID” in the media, formed social-media based support groups which have spearheaded advocacy efforts. Surveys conducted by such advocacy groups suggest that a heterogeneous, multisystemic constellation of symptoms and complications are experienced by these individuals,^14,15^ which has prompted more recent attention to post-COVID syndrome in the medical literature^16-19^ and is consistent with our present findings.

The only longitudinal study of putatively recovered COVID-19 cases found that 98% of 201 participants were still experiencing symptoms four months post illness onset, including fatigue, myalgias, headaches, cardiorespiratory and gastrointestinal symptoms.^20^ Consistent with these symptoms, multi-organ magnetic resonance imaging (MRI) detected impairment in one or more organs, including heart, lungs, kidneys, liver, pancreas, and spleen in 70% of the sample. Few of these participants had any pre-existing health conditions and only 18% were hospitalized during acute illness. While it remains unknown whether some or all of those with post-COVID syndrome will eventually return to their pre-COVID level of health and functioning, post-COVID morbidity is likely to place a substantial burden on healthcare systems that are already depleted by their response to acute COVID-19.

Notably, all but one of the post-COVID syndrome participants in this study were female. While mortality from COVID-19 is sexually dimorphic, with males accounting for approximately 70% of deaths,^21^ the present findings suggest that females are more prone to a prolonged course of illness and/or post-COVID-19 complications, similar to the preprint findings from the Mayo Clinic.^20^ Innate and immune responses are stronger in females than in males.^22^ It is plausible that the more active female immune system may be an advantage in clearing the virus, but this could be a double-edged sword if the female immune activation produces a lingering syndrome. This sexually dimorphic immune response is further supported by a recent study which found that 12.5% of males but only 2.6% of females with severe COVID-19 had neutralizing immunoglobulin G (IgG) autoantibodies against interferons which could explain some of the increased male mortality.^23^

## Limitations

To our knowledge, this is the first report of post-COVID syndrome in young adults and the first to focus exclusively on non-hospitalized cases. However, this study is not without limitations. The sample of positive students is small, although consistent with the positivity rates on campus.^24^ Additionally, the lack of age and racial and ethnic diversity in the sample may limit the generalizability of our findings to more diverse university populations.

Given the low specificity rates of PCR testing, we cannot say with confidence that all students who tested negative for COVID-19 and were never clinician diagnosed did not have the illness. As asymptomatic infections are well documented,^25,26^ it’s possible that a few of our unaffected students could have also been infected, lowering the proportion with persistent symptoms. However, all students who returned to campus were required to get tested prior to their arrival and the university has been randomly testing up to 1,000 students a week. In our sample, 14% of those who were asymptomatic and randomly tested reported a positive result, which reduces the likelihood that a significant number of asymptomatic students are represented in this sample. Still, even if all participants had been infected, a persistent disabling syndrome minimally affected 15% of the sample. Finally, this study relies on retrospective self-report data and longitudinal studies are needed to provide a more nuanced understanding of the long-term sequelae of post-COVID syndrome.

## Conclusion

Our results suggest that young adults are experiencing protracted symptoms and complications post-COVID. There is overwhelming anecdotal evidence that some who contract COVID-19 do not recover within 2-3 weeks; however, the proportion of those who experience post-COVID syndrome at a population level remains unknown. Large-scale population-based studies in larger, more diverse samples are essential to discerning the magnitude and characterization of post-COVID syndrome. Until such studies are executed, we urge the medical and scientific community to consider young adults vulnerable to post-COVID syndrome and to closely monitor those who contract COVID-19 for lingering viral effects.

## Data Availability

Data is available by request. Please request via

## Conflict of Interest

NG is the Founder of the Pulmonary Wellness Foundation, under which he opened the Pulmonary Wellness Foundation’s COVID Rehabilitation & Recovery Clinic. He also serves as a consultant for PulmonX. All other authors declare no conflicts of interest.

## Acknowledgements

Thank you to the following students for their assistance with data collection and Qualtrics programming: Jenna Kilian, Julie Mitchell, Piper Sereno, Madeline Scherer, Maya Quale, Macy Newburg, and Patricia Sabal.

## Notes

**Funding:** This work was supported by NIMH R01MH110418 (DM)

### Competing Interest Statement

Dr. Greenspan serves as a consultant for PulmonX.

### Funding Statement

This work was supported by NIMH R01MH110418 (DM)

### Author Declarations

This study was approved by the University of Dayton Research, Review, and Ethics Committee, a sub-committee of the university Institutional Review Board.

### Summary of Updates

Corrected two typos in author list

